# Novel tools for comparing the architecture of psychopathology between neurogenetic disorders: An application to X- vs. Y-chromosome aneuploidy effects in males

**DOI:** 10.1101/2025.01.10.25320352

**Authors:** Isabella G. Larsen, Siyuan Liu, Lukas Schaffer, Srishti Rau, Tiffany Ajumobi, Bridget W. Mahony, Allysa Warling, Ethan T. Whitman, Ajay Nadig, Cassidy McDermott, Anastasia Xenophontos, Kathleen Wilson, Liv S. Clasen, Erin N. Torres, Jonathan D. Blumenthal, Dani S. Bassett, Armin Raznahan

## Abstract

**Background:** Psychiatric symptoms are typically highly inter-correlated at the group level. Collectively, these correlations define the architecture of psychopathology—informing taxonomic and mechanistic models in psychiatry. However, to date, it remains unclear if this architecture differs between etiologically distinct subgroups, despite the core relevance of this understanding for personalized medicine. Here, we introduce a new analytic pipeline to probe group differences in the psychopathology architecture—demonstrated through comparison of two distinct neurogenetic disorders.

**Methods:** We use a large questionnaire battery in 300 individuals aged 5-25 years (*n* = 102 XXY/KS, *n* = 64 XYY, *n* = 134 age-matched XY) to characterize the structure of correlations among 53 diverse measures of psychopathology in XXY/KS and XYY syndrome—enabling us to compare the effects of X- vs. Y-chromosome dosage on the architecture of psychopathology at multiple, distinctly informative levels.

**Results:** Behavior correlation matrices describe the architecture of psychopathology in each syndrome. Comparison of matrix row averages reveals that autism-related features and externalizing symptoms are most differentially coupled to other aspects of psychopathology in XXY/KS vs. XYY. Clustering the difference between matrices captures coordinated group differences in pairwise coupling between measures of psychopathology: XXY/KS increases coherence among externalizing, internalizing, and autism-related features, while XYY syndrome shows greater coherence in dissociality and early neurodevelopmental impairment.

**Conclusions:** These methods offer new insights into X- and Y-chromosome dosage effects on behavior, and our shared code can now be applied to other clinical groups of interest—helping to hone mechanistic models and inform the tailoring of care.

## INTRODUCTION

It is well established that psychiatric features, such as the presence or absence of individual symptoms or the severity of continuously measured traits, are often highly intercorrelated across individuals (Borsboom & Cramer, 2013; Mahony et al., 2023). Collectively, these correlations capture the architecture of psychopathology, and therefore inform psychiatric taxonomy, mechanistic models, and the optimization of measurement methods in psychiatric assessment.

Recent years have seen increased interest in applying network models to describe and analyze observed correlations between different aspects of psychopathology (Borsboom & Cramer, 2013; Fried & Cramer, 2017; Fried et al., 2017). Borrowing tools from neuroscience and social psychology, the network approach to psychopathology views mental health as a complex web of individual symptoms that interact to generate broader patterns of psychopathology (Borsboom & Cramer, 2013; Fried et al., 2017; Morvan et al., 2020). Network models can depict the architecture of psychopathology in informative ways, by identifying which symptoms are most strongly coupled to all others or discovering sets of symptoms that cluster into coherent modules, for example (Borsboom & Cramer, 2013; Roefs et al., 2022). These insights not only help to refine taxonomic and causal models, but also offer a path to reducing the burden of clinical assessment by identifying core symptoms which efficiently proxy the severity of many others. While network approaches to psychopathology have proliferated in the past decade, they have yet to be used to compare groups with etiologically distinct subtypes of psychopathology (Robinaugh et al., 2020). However, understanding if differing risk factors for psychiatric morbidity yield different architectures of psychopathology is of crucial theoretical, methodological, and practical importance.

Here, to address this gap in research, we develop and apply a framework for comparing the architecture of psychopathology between cohorts with etiologically distinct subtypes of neuropsychiatric impairment. The current paper introduces this approach through illustrative comparison of two categorically distinct neurogenetic disorders—XXY/Klinefelter syndrome (XXY/KS) and XYY syndrome—but our methods can be generalized to compare any two groups of interest. We focus here on XXY/KS and XYY because these two conditions benefit from availability of unprecedentedly deep phenotypic dimensional measures in large samples (Schaffer et al., 2024), which are well-suited to network-based analyses (Raznahan et al., 2023). Also, these sex chromosome aneuploidies (SCAs) are collectively common [combined prevalence ~1/500 males (Nielsen & Wohlert, 1991; Sánchez et al., 2023)], and some of the most penetrant gene dosage disorders for psychiatric outcomes (Sánchez et al., 2023; Vaez et al., 2023). Finally, the comparison of XXY/KS and XYY directly contrasts X- vs. Y-chromosome dosage effects on human behavior, which are of interest to studying normative sex differences in populations beyond SCAs (Green et al., 2019).

The “Genetic-first” approach adopted by the current study has become increasingly popular in psychiatry because genetically defined disorders such as copy number variants (CNVs) and aneuploidies offer a much-needed objective foothold into the complex biology of psychiatric disorders (Raznahan et al., 2022; Sanders et al., 2019). Gene dosage disorders exert large effect size increases in risk or psychopathology that impact diverse transdiagnostic domains and can vary in profile between different CNVs and aneuploidies (Lee et al., 2022; Rau et al., 2021; Raznahan et al., 2022). Observed differences in the profile of average symptom severity between genetically defined groups opens the door to personalized care and helps to fractionate pathways of biological risk for psychopathology. However, genetically defined groups have yet to be compared for the patterns of within-group correlations between different dimensions of psychopathology—a problem that is ripe for treatment using network approaches. If inter-symptom correlations differ between gene dosage disorder groups, then this would indicate genetic control on the coherence of psychopathology and point towards the need for distinct causal models as well as targets for clinical assessment and intervention as a function of genotype. Conversely, establishing equivalence of symptom correlation networks between genetic subgroups supports broader use of common measurement methods and targets for behavioral intervention.

In the current study, we use pairwise correlation matrices encompassing 53 diverse scales of psychopathology for both XXY/KS and XYY syndrome to compare X- vs. Y-chromosome effects on the architecture of psychopathology at 4 distinctly informative levels: (i) the strength of each scale’s correlation with all others; (ii) the profile of each scale’s correlation with all others; (iii) the overall strength of coupling between all scales; and (iv) the strength of pairwise coupling between distinct subsets of scales—capturing sets of clinical features which are more or less coherent in one group than the other. Our methods and findings improve understanding of sex chromosome dosage effects on human behavior while introducing a new suite of analytic methods and shared code that can now be used by other researchers to compare the architecture of psychopathology between any two populations of interest.

## METHODS

### Participants

Our dataset includes 300 individuals from four groups: XXY/KS (*n* = 102), XYY syndrome (*n* = 64), and two groups of typically developing XY individuals age-matched to each of these sex chromosome aneuploidy (SCA) groups (*n* = 74 age-matched to the XXY group and *n* = 60 age-matched to the XYY group; **Table 1**). To be included in this study, individuals with XXY/KS or XYY must have non-mosaic karyotypes and fall between the ages of five and twenty-five years. Individuals with XXY/KS or XYY who were able to provide a blood sample had cytogenetically confirmed non-mosaic karyotypes (based on a minimum of 50 metaphase spreads; Quest Diagnostics, Nichols Institute Chantilly, VA, USA), and those who were unable to provide a blood sample had non-mosaic karyotypes confirmed using reports of genetic testing from their community provider.

**Table 1.**
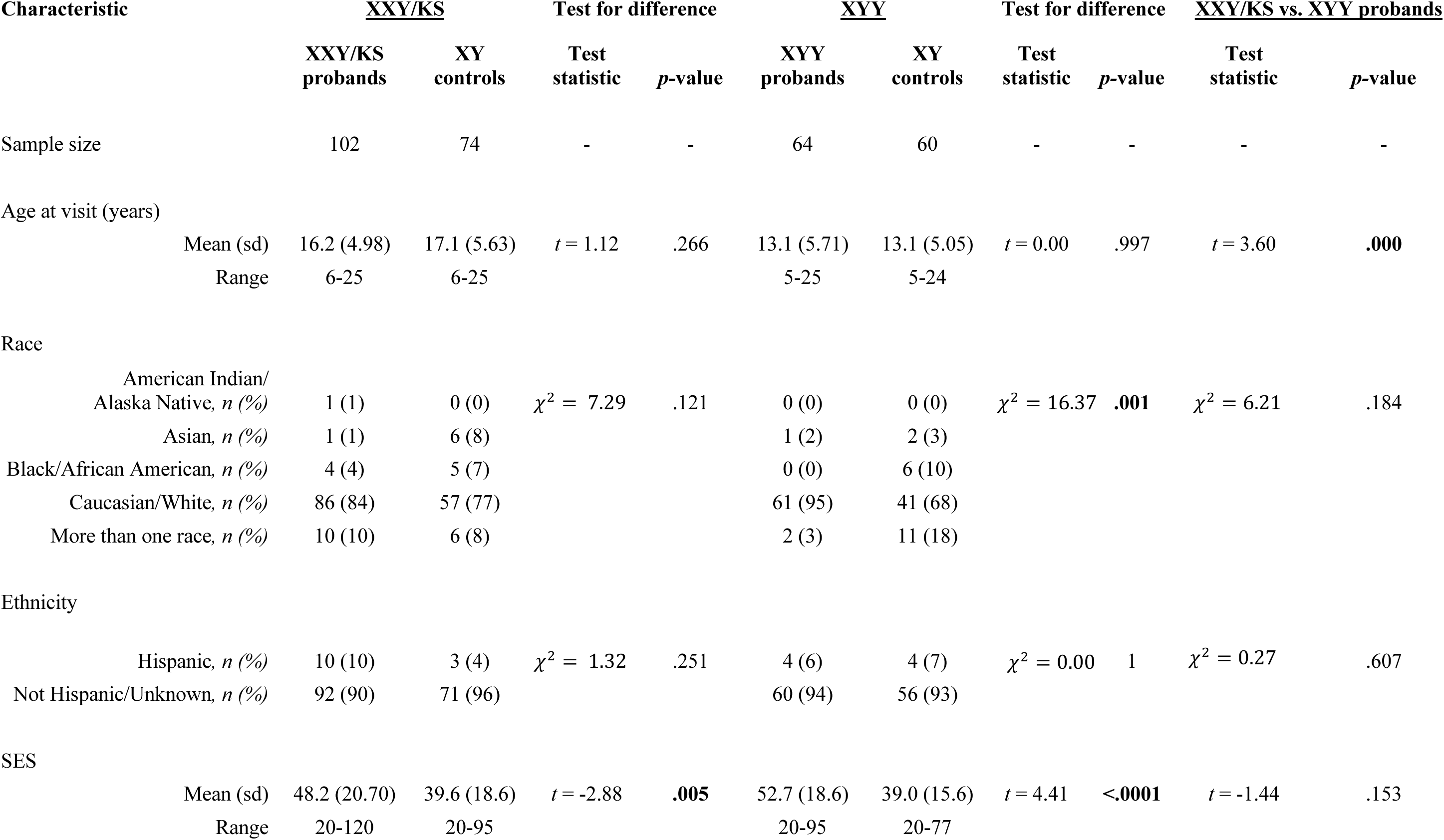
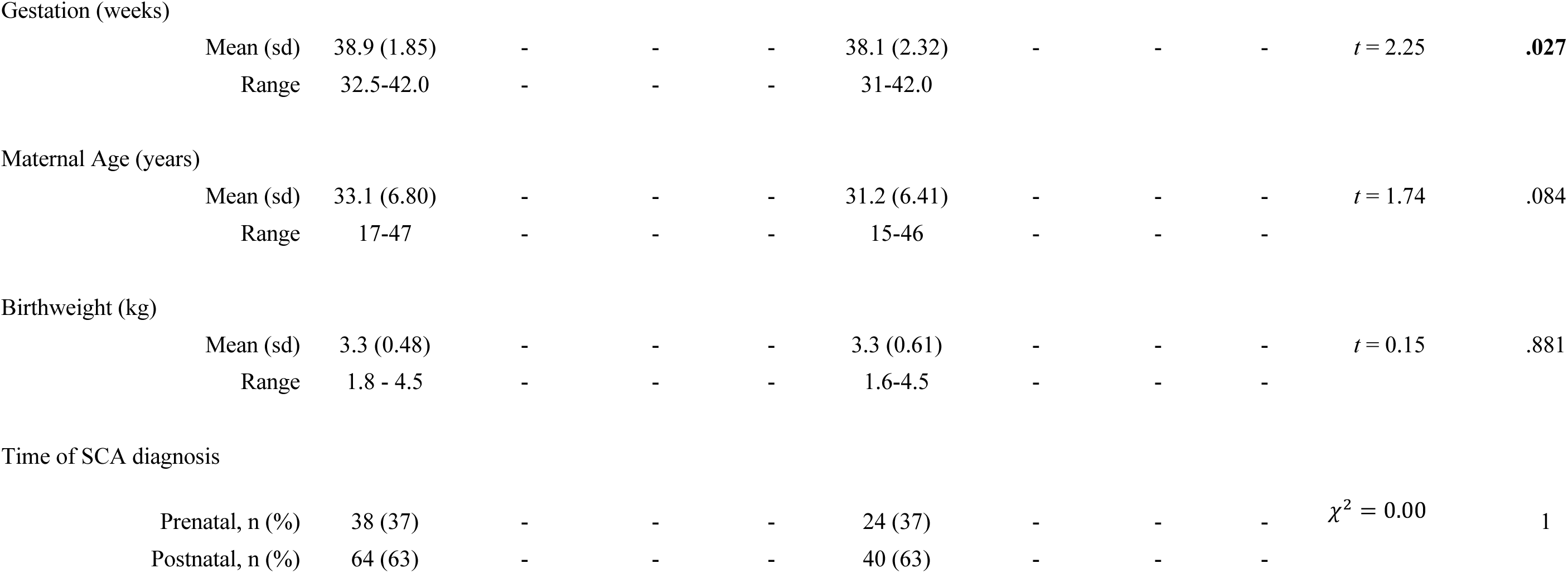
Participant characteristics.

The exclusion criteria for the typically developing XY group included the presence of a psychiatric disorder in the participant or of a severe psychiatric disorder in any of their first-degree relatives (as diagnosed prior to subject study enrollment or identified during the study as part of a standardized interview). All study visits were conducted at the NIH Clinical Center, Bethesda, MD, USA. Participants provided written informed consent (adult participants and parents of minor participants) or written assent (children) prior to completing study procedures. This research was approved by the National Institute of Mental Health Institutional Review Board (NCT00001246).

### Phenotypic Measures

#### Questionnaire-Based Measures of Psychopathology and Behavior

Data from nine self- and parent-report instruments were used in this analysis to provide broad and deep phenotypic information related to psychopathology and behavior. These nine instruments yield 53 distinct dimensional measures (scales) of psychopathology that cut across dimensions of psychosocial and behavioral functioning to allow a fine-grained transdiagnostic characterization of mental health (**Table S1**). The Child Behavior Checklist (CBCL; (Achenbach & Edelbrock, 1983)) and the Strengths and Difficulties Questionnaire (SDQ; (Goodman, 1997)) provide multidimensional descriptions of psychopathology and functioning. Features of Autism Spectrum Disorder (ASD) are characterized by the Social Responsiveness Scale, Second Edition (SRS-2; (Constantino & Gruber, 2012)) and the Social Communication Questionnaire, SCQ; (Rutter et al., 2003)). Diverse other scales capture domains of motor coordination (Developmental Coordination Disorder Questionnaire, DCDQ; (Wilson et al., 2009)), attention-deficit/hyperactivity features (Conners-3 (Conners et al., 2008)), irritability (Affective Reactivity Index, ARI; (Stringaris et al., 2012)), conduct problems and dissociality (Antisocial Process Screening Device, APSD; (Vitacco et al., 2003)), and aggression (Children’s Scale of Hostility and Aggression: Reactive/Proactive, C-SHARP; (Farmer & Aman, 2009), or the Adult Scale of Hostility and Aggression: Reactive/Proactive, A-SHARP (Matlock & Aman, 2011)).

#### Data Preparation and Standardization of Scores

For both SCA groups, individual-level scores for all 53 scales were standardized against those in the corresponding age-matched typically developing XY group as previously described (Raznahan et al., 2023; Schaffer et al., 2022). This procedure ensures that all scores are on a harmonized scale even though many of the originating instruments lack norms, and those with norms vary in their reference populations. Individual-level scores in the SCA groups were scaled in comparison to their XY counterpart using either (1) the observed mean and standard deviation of that score in the XY comparison group (in the absence of significant age-by-group interactions for the given instrument), or (2) scaled residuals for SCA individuals relative to a general linear model in the XY group with age as a linear predictor (given significant age-by-group interactions). To ensure consistent scoring polarity (where higher scores indicate greater impairment), we inverted scores for one subscale and one instrument (prosocial behavior on the SDQ and all scales on the DCDQ). Because the included questionnaires had varying applicable age ranges, the available sample size varied across scales, but all instruments had ratings for at least 108 of 166 total individuals across SCA groups (mean completeness proportion = 0.83, proportion range = 0.65 - 0.98, **Table S1**) and the mean age of participants for each instrument was matched between SCAs and their respective XY controls for both XXY/KS and XYY (**Table S1**).

### Statistical Analysis

Descriptive statistics included proportions for categorical variables and means for continuous variables. Inferential statistics on inter-scale correlation networks used parametric and permutation-based non-parametric tests as detailed under each Network Analysis subsection below. Given that all participants were gonadally male, gonadal sex was not controlled for in analyses. All statistical analyses and data visualizations were conducted using the R language for statistical computing (R Core Team, 2023).

#### Computing the effect size of XXY/KS and XYY on each scale

The effect size of each SCA on each of the 53 scales was estimated using linear models as previously described (Raznahan et al., 2023; Schaffer et al., 2022). Specifically, in a linear model with each scale’s standardized scores across the SCA group and their corresponding XY control group as the dependent variable, the coefficient for the binary variable of group estimates the standardized effect size of the SCA on each scale.

#### Constructing scale correlation networks in each group

We computed Pearson correlation coefficients for the pairwise relationships between all scales across individuals in each SCA group (using scores normalized against XY controls as detailed above)—resulting in one square, symmetric 53*53 correlation matrix for XXY/KS and another for XYY syndrome. These matrices can each also be conceptualized as signed, weighted and unthresholded networks consisting of 53 nodes (one for each measured scale) connected by edges (correlation coefficients capturing the strength of coupling between each unique pair of scales). We report raw Pearson correlation coefficients for all within-group descriptions of inter-scale relationships to ensure ease of interpretability and consistency with prior work (Raznahan et al., 2023). Pearson correlation coefficients were Fisher’s *Z*-transformed for all other analyses given the greater suitability of these transformed scores for parametric inferential statistics (Meng et al., 1992).

#### Measuring and comparing the overall coupling of each individual scale to all others

For each syndrome network, mean weighted nodal degrees capture the overall strength of connectivity (coupling) between each node (scale) and all others. We calculated nodal degree for each of the 53 measured scales in each syndrome as the mean Fisher’s *Z*-transformed Pearson’s *r* across each row of the syndrome’s 53*53 pairwise correlation matrix (**Figure 1B**). We used correlation and Deming-regression to compare these two vectors of 53 nodal degree values between syndromes. The Pearson correlation between the 53 nodal values captures the overall congruence of these values between syndromes. Deming regression (which we implemented using the R package *SimplyAgree* (Caldwell, 2022)) provides a formalized framework for testing if the relationship between nodal degree values in XXY/KS and XYY deviates from identity. This relationship can be visualized as a scatterplot where each point is a scale, the two axes are nodal degree in XXY/KS and XYY and the line x=y defines where points will fall if they have identical degrees in both syndromes (**Figure 1C**). Deming regression defines the orthogonal least squares fit line for this scatterplot, which—unlike traditional least square regression—provides the same fit line regardless of which syndrome’s degree is the dependent versus independent variable (Adcock, 1878; Pallavi et al., 2022).

**Figure 1.**
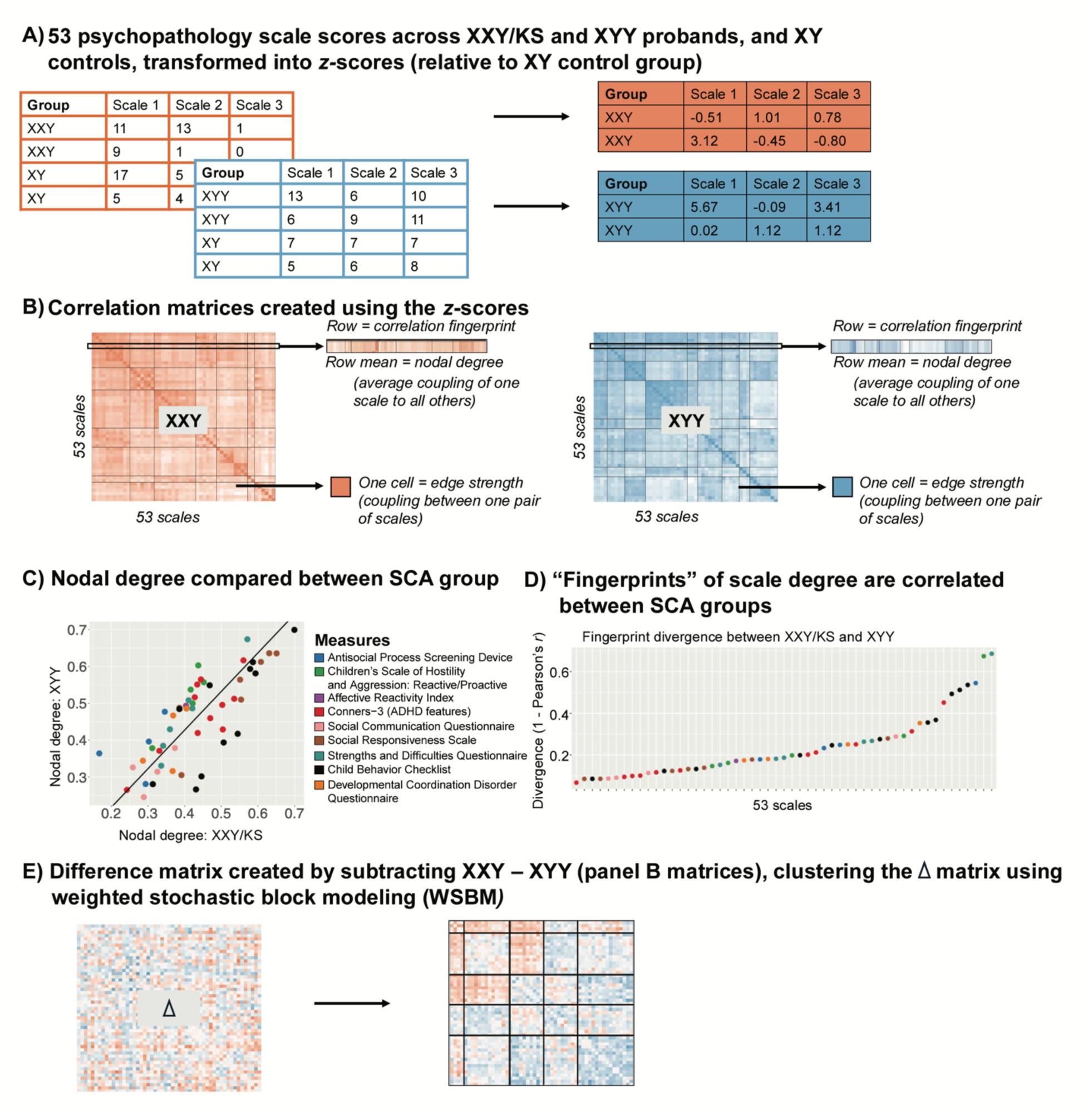
Schematic Overviewing Analytic Workflow. **(A)** Raw scores for each of 53 scales are scaled in each SCA group using the mean and standard deviation in their corresponding XY control group. **(B)** Cross-individual correlations between each unique pair of scales yield a 53*53 square, symmetric correlation matrix for both the XXY/KS and XYY group. This matrix can be conceptualized as a signed, weighted and unthresholded network, where each node is a scale and each edge captures the strength and direction of correlation between scales. A single row of these matrices captures the “fingerprint” of a single scale’s correlation with all other scales. Row averages capture each scale’s “nodal degree,” indexing the overall strength of its correlation with all other scales. (**C**) Correlating nodal degree values between groups (across scales) quantifies the overall similarity of each scale’s connectivity with all others in XXY/KS and XYY and highlights scales that differ most in the overall strength of their coupling with other scales between groups. (**D**) Correlating fingerprints between groups identifies scales that are most divergent in the specific profile of their coupling with all others between XXY/KS and XYY. (**E**) Subtracting the two matrices in panel (B) yields a single 53*53 square symmetric matrix of the differences in correlation between each unique pair of scales in XXY/KS vs. XYY. Clustering this difference matrix identifies sets of edges that show coordinated differences in strength between groups (i.e., sets of scales that are more or less coherent in one group vs. the other).

To test for scales with a statistically significant difference in nodal degree between syndromes we compared the observed difference in nodal degree for each scale to a distribution of null degree differences for 10,000 random permutations of group membership across all 166 individuals with SCA. The empirical *p*-value for group differences in each scale was defined as the percent of absolute permuted values exceeding the observed value. These *p*-values were Bonferroni-corrected for 53 comparisons across scales (*p* < .05/53, or *p* < .0009).

#### Characterizing and comparing the fingerprint of each scale’s connectivity to all others

Nodal degree captures the overall magnitude of a scale’s connectivity with all other measured scales. In contrast, the full vector of 53 correlations that is averaged to compute nodal degree captures the profile or “fingerprint” of a scale’s connectivity with all other scales (**Figure 1B**). We computed a “fingerprint divergence score” to estimate the difference in each scale’s connectivity fingerprint between XXY/KS and XYY syndrome. Specifically, we computed the Pearson correlation between syndromes for the vector of 53 behavioral correlation values associated with each scale and converted them to a “fingerprint divergence score” (1 – correlation) (**Figure 1D**). Higher scores indicate greater differences in scale connectivity values and lower values indicate more similar connectivity fingerprints between syndromes. We tested for deviation of these fingerprint divergence scores from 0 by comparing the observed fingerprint divergence score to a null distribution across 10,000 permutations of group membership. The resulting *p-*value is the proportion of times the absolute divergence score in the null distribution exceeded the observed absolute divergence score, divided by the number of permutations. For scales with significant fingerprint divergence between XXY/KS and XYY syndrome, we used a scatterplot, Deming regression, and permutations (as described above) to identify which specific inter-scale relationships were driving this divergence between syndromes. Finally, we used correlation analysis across all 53 scales to test if fingerprint divergence between XXY/KS and XYY is related to the absolute difference in effect size of XXY/KS and XYY on each scale (**Table S1**; **Figure 2D**).

**Figure 2:**
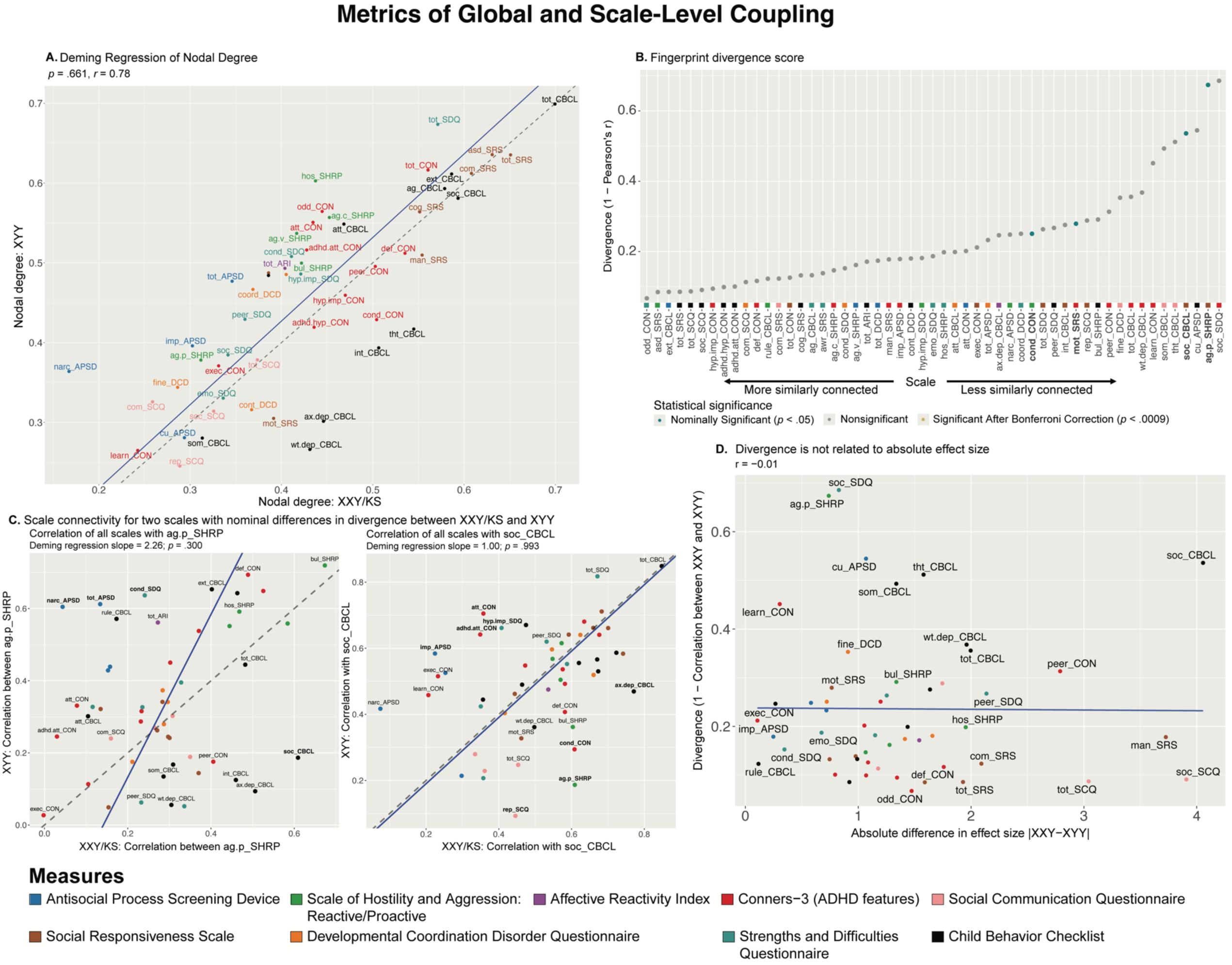
Comparing XXY/KS and XYY for the magnitude and profile of psychopathology connectivity at the level of individual behavioral scales. **(A)** Scatterplot showing the relationship between nodal degree (magnitude of a scale’s overall connectivity) for 53 different behavioral scales in XXY/KS vs. XYY. The identity y=x line is dashed gray. The fit line from Deming regression is solid blue. Scales are colored by the domain of psychopathology they measure. **(B)** Dot plot showing the divergence between nodal edge profiles (the profile of a scale’s connectivity with all others) in XXY/KS vs. XYY for 53 different behavioral scales. Dot color and bold text indicate scales with statistically significant divergence scores. **(C)** Scatterplots showing the correlation between each scale and two of the nominally significant scales from panel (B) in each group, ag.p_SHRP and soc_CBCL. Bolded scale labels indicate scales with nominally significant (*p* < .05) differences in correlation between groups. The Deming regression fit line is solid blue and the identity y=x line is dashed gray. Scales are colored by the domain of psychopathology they measure. **(D)** Scatterplot showing the relationship between the absolute difference in effect size of XXY/KS vs. XYY on the mean score of each behavioral scale (x-axis) and the difference in connectivity profile of each scale with all others in XXY/KS vs. XYY (y-axis).

#### Fine-grained comparison of coupling between individual scale pairs

The analyses above compare the architecture of psychopathology between syndromes at the level of individual scales. We next sought to compare syndromes for the full distribution of all 1,378 unique pairwise correlations between scales. We used Fisher’s Z-transformed correlations for these analyses and compared the distribution of these values using three complimentary tests. First, the means of these distributions were compared as the absolute difference between XXY/KS and XYY (XXY/KS – XYY, or Δ*z*). Given the large number of observations, we assessed statistical significance of the observed absolute difference in mean *z* against a distribution of null Δ*z* values from repeated analyses after 10,000 permutations of karyotype group membership. Second, we compared the shape of these two distributions using a two-sample Kolmogorov-Smirnov (KS) test, using the *ks.test()* function from the R package *stats*. We report the test statistic, *D* (a measure of the maximum distance between the two empirical cumulative distribution functions (Berger & Zhou)) and the *p*-value calculated from the null distribution resulting from our permutation with 10,000 iterations. Third, we compared the weight of the tails of the edge strength distribution between groups using the *kurtosis()* function from the R package *moments* (Lukasz Komsta & Novomestky, 2022).

#### Defining sets of pairwise relationships between scales that show coordinated differences between syndromes

We hypothesized that meaningful differences in the architecture of psychopathology between groups should manifest as coordinated differences of inter-scale correlation for sets of related scales. For example, because our battery of 53 scales includes more than one measure for constructs of inattention and low mood, an altered coupling of the two constructs between syndromes should manifest as a coordinated difference of inter-scale correlations amongst the multiple scales in our battery that relate to these two constructs.

To characterize coordinated group difference in inter-scale connectivity, we first subtracted the Fisher’s Z-transformed inter-scale correlation matrices for XXY/KS and XYY to generate a single a 53*53 delta matrix (Δ matrix) where each cell represents the difference between syndromes in the pairwise correlation for a single pair of scales (**Figure 1E**). In this matrix, negative Δ*z* values indicate that the pair of scales is more strongly correlated in XYY, while positive Δ*z* values indicate that the pair of scales is more strongly correlated in XXY/KS. This Δ matrix was then clustered using Weighted Stochastic Block Modelling (WSBM; with the function *BM_gaussian()* from the R package *blockmodels* (Leger et al., 2021)) to define sets of scale pairs that show coordinated differences in correlation between syndromes (**Figure 1E**). As a complement to this analytic view, we also ran supplementary analyses where WSBM was applied to the behavioral correlation matrix of each syndrome separately in order to identify scales that differed in cluster assignment between syndromes (**Figure S2**).

WSBM is a generative algorithm that has proved highly effective in clustering behavioral correlation matrices because it accounts for both the connectivity between scales and the connectivity across clusters, and it can detect both assortative and non-assortative (e.g., core-periphery; (Betzel et al., 2018)) classes of cluster solution. The optimal number of clusters was selected from the WSBM by taking the clustering solution that maximized the integrated classification likelihood (ICL; (Biernacki et al., 2000)). Clusters were given descriptive names based on their content and following prior work (Raznahan et al., 2023). Correlation matrices were visualized using a heatmap created with the R package *superheat* (Barter & Yu, 2023) and a network visualization of the WSBM clustered Δ matrix was created using the R package *igraph* (Csardi & Nepusz, 2006).

#### Applying WSBM to the Δ *matrix* groups nodes into clusters

The sets of edges that fall within individual clusters and between each unique pair of clusters are referred to as blocks. Blocks represent sets of scale-pairs that show similarly altered inter-scale correlations in XXY/KS vs. XYY. The average value of all correlations within a block estimates the overall magnitude and direction of differences in correlation between syndromes. We use a previously developed method (Mahony et al., 2023) to identify blocks with significantly extreme changes in correlation between syndromes. The defined test statistic for each block was the mean edge strength of all edges after excluding the diagonal. We compared each block’s observed mean edge value to a null distribution of mean edge values from 10,000 permutations of SCA group membership. For each permutation, SCA karyotype was randomized across all 166 SCA individuals and scale correlation matrices were re-generated for the resulting groups. These matrices were then subtracted from each other, and the edge values in this null Δ matrix were used to compute null mean edge strength values for the sets of edges defining each block in our observed WSBM solution. As such, the mean observed edge weight for each WSBM block could be compared against a null distribution of mean edge weights for the same edges from permutation. The empirical *p*-value for group differences in each block was defined as the percent of absolute permuted values exceeding the observed value. These *p*-values were Bonferroni-adjusted for multiple comparisons across unique blocks.

## RESULTS

### Participant Characteristics

Participant characteristics are described in **Table 1**. The full sample of 300 individuals included 166 individuals with SCA (102 XXY/KS, 64 XYY syndrome) and their age-matched XY control groups for each SCA subgroup (74 for XXY/KS, and 60 for XYY syndrome). The two SCA groups differed in mean age at the time of the visit (XXY/KS = 16.2 years, XYY = 13.1 years, *p* = .001)**—**hence, each group is paired with an independent contemporaneously recruited group of age-matched XY controls. XXY/KS and XYY groups also showed a statistically significant, but small (~1 day) difference in mean gestational age (XXY/KS = 38.9 weeks, XYY = 38.1 weeks, *p* = .027). There were no significant differences between the XXY/KS cohort and the XYY cohort in race, ethnicity, socioeconomic status (SES), maternal age, birthweight, or time of SCA diagnosis (prenatal vs. postnatal).

### Comparing the strength of each scale’s connectivity with all others

A group-level behavioral correlation matrix makes it possible to compute each behavioral scale’s correlation with all others (a “global coupling score,” i.e., the nodal degree value for each scale). Global coupling scores capture the overall strength of within-group correlation between any single scale and all other measured scales. We took several complementary approaches to compare these scale-level global coupling scores between XXY/KS and XYY syndrome.

First, we found that scale-level global coupling scores in XXY/KS and XYY are highly correlated with each other (Pearson correlation across all 53 scales, *r* = 0.78, **Fig 2A**). For both syndromes, the highest global coupling scores were seen for summary measures of overall psychopathology and ASD-related features, whereas the lowest global coupling scores were seen for measures of learning problems, somatic problems and callous-unemotional traits. Second, Deming regression revealed that the fit line describing the relationship between scale-level global coupling scores in XXY/KS and XYY is statistically indistinguishable from the y=x identity line (*p*-values for tests of slope vs. 1 and intercept vs. 0, *p* = .661 and *p* = .875, respectively). Third, the largest residuals from the Deming regression fit line (**Table S1**) were seen for measures of hostility and antisocial symptoms (higher degree in XYY than XXY/KS) as well as those of internalizing disorder symptoms (higher degree in XXY/KS than XYY). However, no single scale showed a statistically significant difference in global coupling between XXY/KS and XYY that survived correction for multiple comparisons after permutation-based testing. Taken together, these results establish that XXY/KS and XYY are broadly similar in the extent to which any given aspect of psychopathology is, on average, correlated to others.

### Comparing the profile of each scale’s connectivity with all others

A given scale may be similarly strongly coupled to all other scales in two different clinical groups but differ between groups in the specific profile—or connectivity fingerprint—of its correlations with all other scales. A scale could have a different connectivity fingerprint between two groups if groups differ in the causal relationships between that scale and all others, or in the psychometric properties of the instruments that measure the scale in question.

Despite scales showing highly similar global coupling scores between XXY/KS and XYY (**Figure 2A**), we found that scales varied widely in the magnitude of their connectivity fingerprint divergence (See Methods) between the two syndromes [0.07 ≤ (1 − *r*) ≤ 0.69; **Figure 2B**, **Table S1**]. Measures of proactive aggression (ag.p_SHRP) and social problems (soc_CBCL) showed the largest connectivity fingerprint divergence between XXY/KS and XYY (corresponding to weak correlation values: ag.p_SHRP, *r* = 0.33; soc_CBCL, *r* = 0.46). A total of four measures—all involving social and conduct problems—had connectivity fingerprints divergence scores with nominally significant elevations above 0 (*p* < .05 uncorrected, **Figure 2B**). Comparison of XXY/KS and XYY for each scale’s coupling with the C-SHRP measure of proactive aggression (ag.p_SHRP) revealed that proactive aggression was more strongly coupled with social problems in XXY/KS vs. XYY, but more strongly coupled with multiple measures of externalizing behavior in XYY vs. XXY/KS (**Figure 2C** left panel). A similar analysis for the CBCL measure of social problems revealed that this aspect of psychopathology was more strongly coupled with measures of externalizing and internalizing psychopathology in XXY/KS vs. XYY, but more strongly coupled with multiple measures of inattention and impulsivity in XYY vs. XXY/KS. These differences suggest the need for syndrome-specific measurement methods and/or causal models for social and conduct problems in XXY/KS vs. XY.

Fingerprint divergence scores were uncorrelated with differences in the effect size of XXY/KS vs. XYY on scale scores (**Figure 2D**, **Table S1**)—indicating that comparing the severity of psychopathology across scales vs. the profile of psychopathology connectivity across syndromes gives non-redundant and complementary information.

### Comparing the full distribution of pairwise coupling between all scales

Patterns of behavioral coupling can also be compared between groups by considering the full distribution of pairwise correlation values between all scales. Comparing these distributions provides a way of testing for group differences in the overall coherence of psychopathology (mean of the edge strength distribution) and prevalence of extremely high or low inter-scale coupling values.

Group comparison revealed that the average edge weight [Fisher’s *Z*-adjusted pairwise Pearson correlation coefficients (*z*)] was not significantly different between XXY/KS and XYY based on a permutation testing (XXY/KS mean *z* = 0.43, XYY mean *z* = 0.46, absolute Δ*z* = 0.03, *permuted p* = .682; **Figure 2**). Notably, both edge distributions were right-skewed. The distribution of edge values appeared more leptokurtotic (fat-tailed) in XYY than XXY/KS, but quantitative analysis did not support this conclusion. Both edge distributions were empirically leptokurtic (kurtosis value > 3), with kurtosis values of 9.84 (XXY/KS) and 7.65 (XYY). A two-sample Kolmogorov-Smirnov (KS) test found no significant difference between the distribution of edge strength in XXY/KS compared to that of the XYY group (*D* = 0.07, *p* = .701). Taken together, these results indicate that while the edge strength appears to differ qualitatively between SCA groups such that XYY displays more polarized patterning of coupling across scales, these differences are not statistically significant.

### Identifying sets of scales with coordinated differences in coupling between groups

Given a behavioral correlation matrix in each of the two groups, clustering the difference between these matrices identifies sets of scales that show coordinated differences in their correlations between groups. Weighted stochastic block modeling (WSBM) of the difference in behavioral correlation matrices (the Δ matrix) between XXY/KS and XYY syndrome grouped all 53 scales into six clusters (Methods, **Figure S1**). Visualizing this cluster solution (**Figure 3A**) and assignment of scales to clusters revealed several patterns of note. First, each WSBM cluster brought together scales (often from different questionnaires) that measured similar constructs. Thus, we observe dissociable difference of behavioral correlations in XXY/KS vs. XYY syndrome for measures of internalizing problems, externalizing problems, ASD-related features, two subsets of early neurodevelopmental differences (socio-motor vs. socio-attentional), and dissociality. These same inter-syndrome differences in the coupling between scales were also evident when comparing the assignment of scales to separate WSBM cluster solutions for behavioral correlation matrices within each individual syndrome (Methods, **Figure S2**).

**Figure 3:**
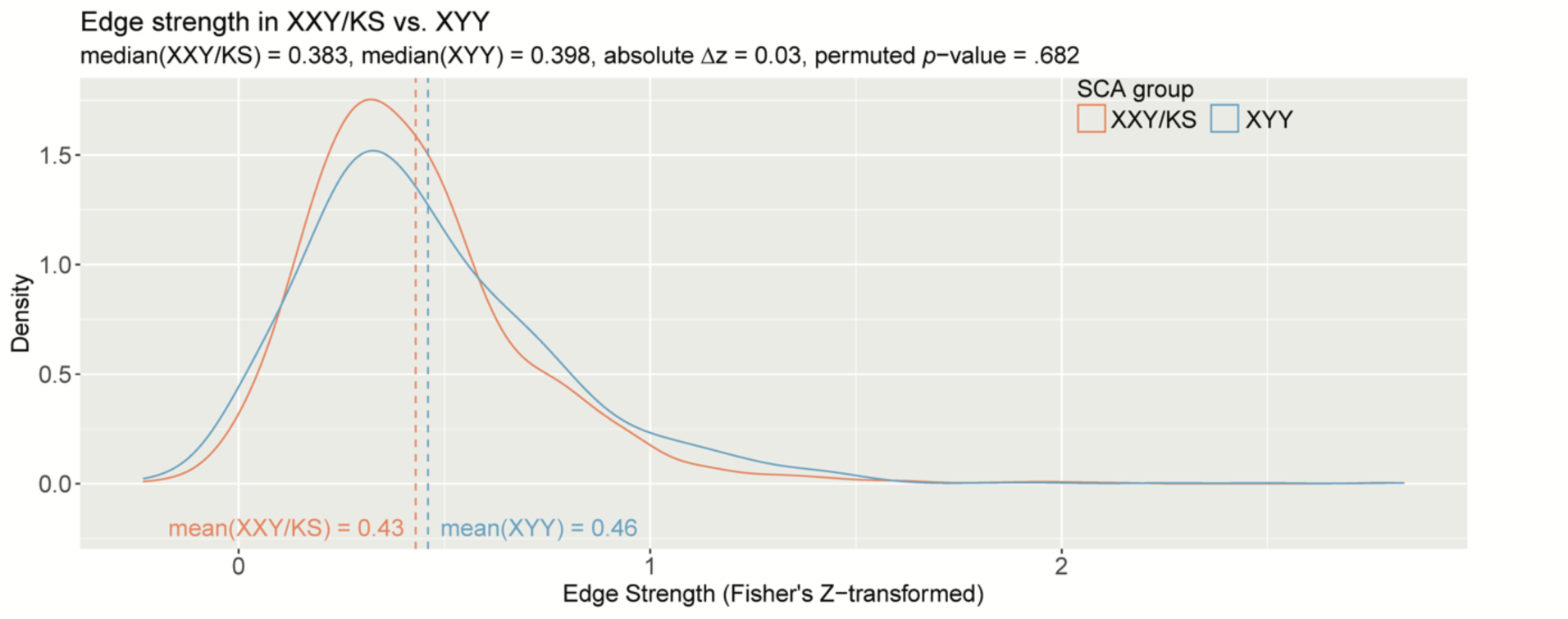
The distribution of all inter-scale correlations in XXY/KS vs. XYY. Density plots show the distribution of Fisher’s *Z*-transformed correlations for all unique pairs of scales in XXY/KS (red) and XYY (blue) groups. Mean edge strength for each group are indicated with dashed vertical lines and specified in inset text, together with the observed difference in mean edge strength (Δ*z*) and the permutation-based *p*-value for this group difference statistic.

Comparing the observed mean correlation values within and between these clusters identified five sets of inter-scale relationships with nominally significant differences (uncorrected *p*<.05) between XXY/KS and XYY syndrome. Within-cluster edges were significantly negative for the cluster of dissociality measures (mean block-wise Δ*z* = −0.23, *p* = .035), indicating that there is greater coherence amongst diverse measures of externalizing symptoms and dissociality in XYY syndrome as compared to XYY/KS. XYY also showed significantly greater coherence than XXY/KS between the two sub-clusters of early neurodevelopmental differences (mean block-wise Δ*z* = −0.22, *p* = .038), indicating more consistency in co-occurrence of early social, motor and attentional impairments in XYY syndrome. Conversely, we observed significantly positive inter-cluster correlation values in XXY/KS vs. XYY for all pairwise relationships amongst three clusters: ASD-related symptoms, internalizing and externalizing problems (ASD-internalizing: mean Δ*z* = 0.21, *p* = .031 / ASD-externalizing: mean block-wise Δ*z* = 0.19, *p* = .037 / internalizing-externalizing: mean Δ*z* = 0.28, *p* = .006). These coordinated group differences in behavioral correlations can also be visualized as a network with each behavioral cluster as a node (**Figure 3B**). This visualization highlights the coordinated “tightening” of correlations amongst ASD, internalizing, and externalizing features in XXY/KS vs. XYY—with internalizing features having the most collectively increased correlation with other features of psychopathology in XXY/KS vs. XYY.

**Figure 4:**
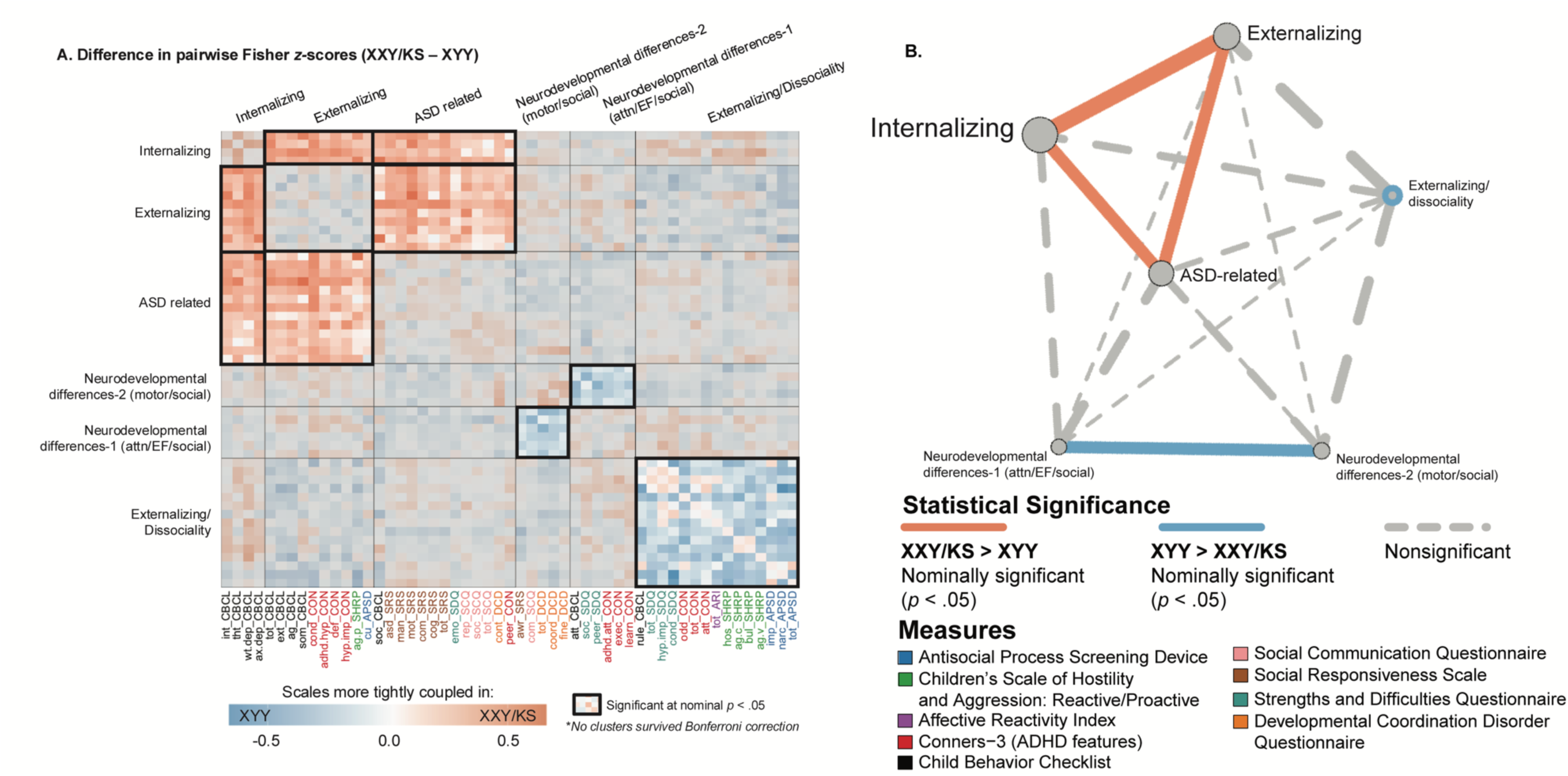
Fine-grained differences between XXY/KS and XYY syndrome in the coupling between different domains of psychopathology. (**A)** Heatmap depicting the weighted stochastic block modeling (WSBM) solution of six clusters (outlined in black) for the delta matrix (Fisher’s *Z*-transformed correlations of XXY/KS – XYY). Scale names are color-coded to the instrument (see **Table S1**). Heatmap cells encode the direction (red: XXY/KS > XYY / blue: XYY > XXY/KS) and magnitude (hue intensity) of group differences in correlation between each unique pair of scales. Blocks with significantly non-zero mean edge strength (nominal *p* < .05) are outlined in bolded black, while blocks within non-significant average group differences in coupling (i.e. mean edge strength statistically indistinguishable from zero) are greyed out. **(B)** Network representation of the WSBM solution. Nodes (circles) represent blocks, and the lines represent edges color-coded by statistical significance. The thickness of the line depicts the edge strength between any two blocks and the node size represents the weighted nodal degree.

## DISCUSSION

Our study introduces a suite of new analytic approaches that test for group differences in the architecture of psychopathology—which we use to compare groups with distinct genetic risk factors for psychopathology. These analyses probe for potential group differences at multiple complementary levels of analyses, each of which carries a different set of conceptual and practical implications. We illustrate each analysis by application to the comparison between two different sex chromosome aneuploidy syndromes—XXY/KS and XYY—with the twofold goal of: (i) determining if and how the architecture of psychopathology differs between these two genetic syndromes; and (ii) providing a worked example so that our shared code can be easily applied to compare correlations between different domains of psychopathology for any two groups of interest. We discuss each of these goals below.

Previous studies have found that XXY/KS and XYY both increase risk for diverse domains of psychopathology, and the relative ranking of different domains by severity of impact is highly conserved between these two SCA groups (Schaffer et al., 2024). At the same time, there are some inter-SCA differences: to the extent that a given domain is impacted in XXY/KS, it tends to be more severely impacted in XYY syndrome, and this disparity is especially marked for social problems (Schaffer et al., 2024). A notable exception to this general rule, however, is symptoms of mood and anxiety, which appear to be similarly elevated in XXY/KS and XYY syndrome (Schaffer et al., 2024). This mixed picture regarding similarities and differences between syndromes in the profile of psychopathology across different domains is echoed in the present study, which focuses instead on comparing syndromes at the level of correlations between different domains of psychopathology. Thus, while we find that the overall strength of a given scale’s correlation with all others is highly conserved between the two SCA groups (highest nodal degree for social and total problems, lowest nodal degree for learning and somatic problems), some scales show substantial group differences in the profile of their correlation with other scales (**Figure 2C**). For example, the coupling of proactive aggression to other measures of psychopathology (its “connectivity fingerprint”) was only weakly correlated between XXY/KS and XYY. Additionally, proactive aggression was more strongly coupled to other scales related to antisocial behaviors and attention problems in XYY, while it was more strongly coupled with internalizing and social problems in XXY/KS. The coupling of social problems to all other scales was also weakly correlated between groups. Social problems were more strongly coupled to features of attention and impulsivity problems in XYY, while they were more strongly coupled to aggression, repetitive behaviors, and internalizing problems in XXY/KS. These findings strongly suggest a different set of causal mechanisms for proactive aggression and social problems in XXY/KS vs. XYY syndrome—highlighting the potential need for genotypically informed methods of clinical assessment and treatment which can now be clarified by targeted comparative studies of aggressive behavior in these two SCA groups.

A mixed picture is also seen for edge-level comparisons of the pairwise correlations between scales in XXY/KS vs. XYY. When all edges (pairwise scale correlations) were considered as a single set, both the mean edge strength and the distribution of edge strengths were statistically indistinguishable between syndromes. However, clustering analysis revealed coherent subsets of edges that collectively differed in strength between XXY/KS and XYY syndrome. Specifically, we observed subtle group differences (surviving nominal *p* < .05 threshold) whereby XXY/KS shows a stronger coherence amongst autism-related, internalizing, and externalizing features, while XYY shows a stronger coherence amongst attentional and motor control neurodevelopmental problems. These findings imply that XXY/KS and XYY must either differ in: the causal relationships between these differentially coupled domains of psychopathology; the relationships between these domains of psychopathology and unmeasured variables; or the construct validity of the measurement instruments used to score symptom severity within these domains of psychopathology. For example, to the extent that some measures capture state- and trait-level behaviors, group differences could reflect differences in temporal co-occurrence. Thus, there is a need for genotypically-tailored mechanistic models and/or measurement approaches for these domains of psychopathology in SCA.

We hope that the application of these methods for comparison of X- vs. Y-chromosome effects motivates future implementation of this pipeline, using our shared code, to other group comparisons of interest in psychiatry. For example, this approach could be applicable to comparisons of normative sex differences within a clinical group, or to comparisons of diagnostic subtypes, such as inattentive versus hyperactive/impulsive subtypes of ADHD. Specifying if and how the architecture of psychopathology differs between groups has diverse theoretical and practical implications. Establishing an equivalent symptom correlation network in two groups provides an empirical basis for using similar approaches of symptom measurement and argues that—at the behavioral level at least—there is unlikely to be a major group difference in the causal relationships between different domains of psychopathology. Conversely, observing a group difference in symptom correlation networks has multiple implications that vary depending on the specific type of group difference observed. First, observing a group difference in the ranking of measured scales by their degree (global coupling score, **Fig 1**, **Fig 2A**) indicates that the relative centrality of different symptom domains to total load of psychopathology is not equivalent between groups. This scenario could carry important practical implications at several different levels including: how to maximize measurement validity in each group (a given symptom dimension could show degree differences between groups because of measurement invariance); how to maximize efficiency of assessment for each group (symptom dimensions with very high degree scores may provide efficient proxies for symptom severity in the network as a whole); and, how to tailor theories regarding potential causal relationships between psychopathology domains in each group (symptom dimensions with very high degree may be core drivers or common outputs of symptom scores across the network as a whole).

Second, observing group differences in the strength of coupling between subsets of psychopathology domains raises a potential need for group-specific measurement, theoretical modeling and intervention around this subset of clinical features. In particular, analysis of fingerprint divergence scores (**Figure 2B,C**) and clustering of scale pairings with different coupling strengths in XXY/KS vs. XYY (**Figure 3**) offer specific testable hypotheses for syndrome differences in the potentially causal relationship between different aspects of psychopathology. For example, the fact that proactive aggression is more strongly coupled to social and affective problems in XXY/KS than XYY, but less strongly coupled to antisocial-related traits, points towards the possibility that aggression is more closely linked to affect regulation issues in XXY/KS, but more closely linked to the core independent axis of dissocial behavior in XYY. This notion proposes that mood features may be more relevant for the genesis and treatment of proactive aggression in XXY/KS than in XYY. These findings further make the case that SCAs cannot be studied as a monolith and instead, separate analyses and comparisons across SCA groups may provide greater nuance and accuracy.

Our results should be interpreted in the context of several study limitations which identify important areas for future work. Although our sample size is large for comparisons of rare neurogenetic disorders and benefits from the large effect sizes associated with such conditions, the absolute number of participants limits our capacity to test pipeline performance over a wide range of input data types and pipeline parameter choices. Future studies in larger sample sizes will be able to refine understanding of pipeline performance and would ideally include the longitudinal data needed to assess temporal stability and more directly test causal hypotheses.

## CONCLUSIONS

Notwithstanding the above limitation and caveats, our study presents a generalizable suite of methods for mapping group differences in the architecture of psychopathology and, by illustrating these methods through application to SCAs, finds subtle yet meaningful differences in this architecture between two distinct genetic risk factors for psychiatric morbidity.

## Data Availability

The datasets used and analyzed during the current study are available from the corresponding author on reasonable request.

## ACKNOWLEDGEMENTS

We thank the patients and their families for participating in this study, as well as the Association for X and Y Chromosome Variations (https://genetic.org) for their assistance with recruitment. We also thank Audrey Thurm, Lisa Joseph, and Cristan Farmer for their roles in data collection.

## ETHICAL STANDARDS

The authors assert that all procedures contributing to this work comply with the ethical standards of the relevant national and institutional committees on human experimentation and with the Helsinki Declaration of 1975, as revised in 2008. This research was approved by the National Institute of Mental Health (NIMH) Institutional Review Board (ClinicalTrials.gov Number NCT 00001246).

