## Supplemental figures and table. for "Novel tools for comparing the architecture of psychopathology between neurogenetic disorders: An application to X- vs. Y-chromosome aneuploidy effects in males"

**SUPPLEMENT**

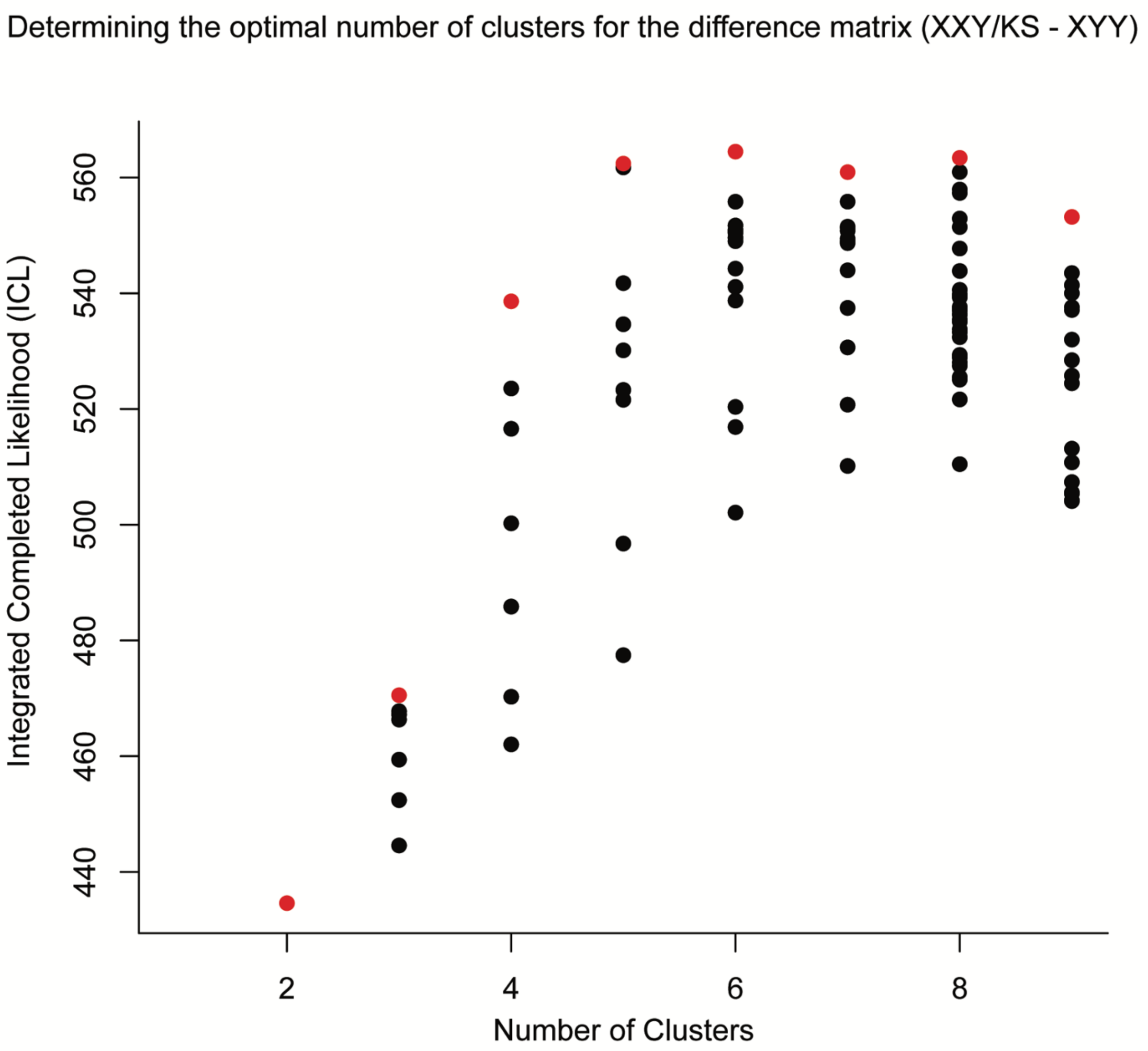


**Figure S1: Optimal cluster determination.** Plot depicting the integrated completed likelihood (ICL) value across iterations of the Weighted Stochastic Block Model. The optimal number of clusters was defined as the number that maximized the ICL value (clusters = 6 for the difference matrix).

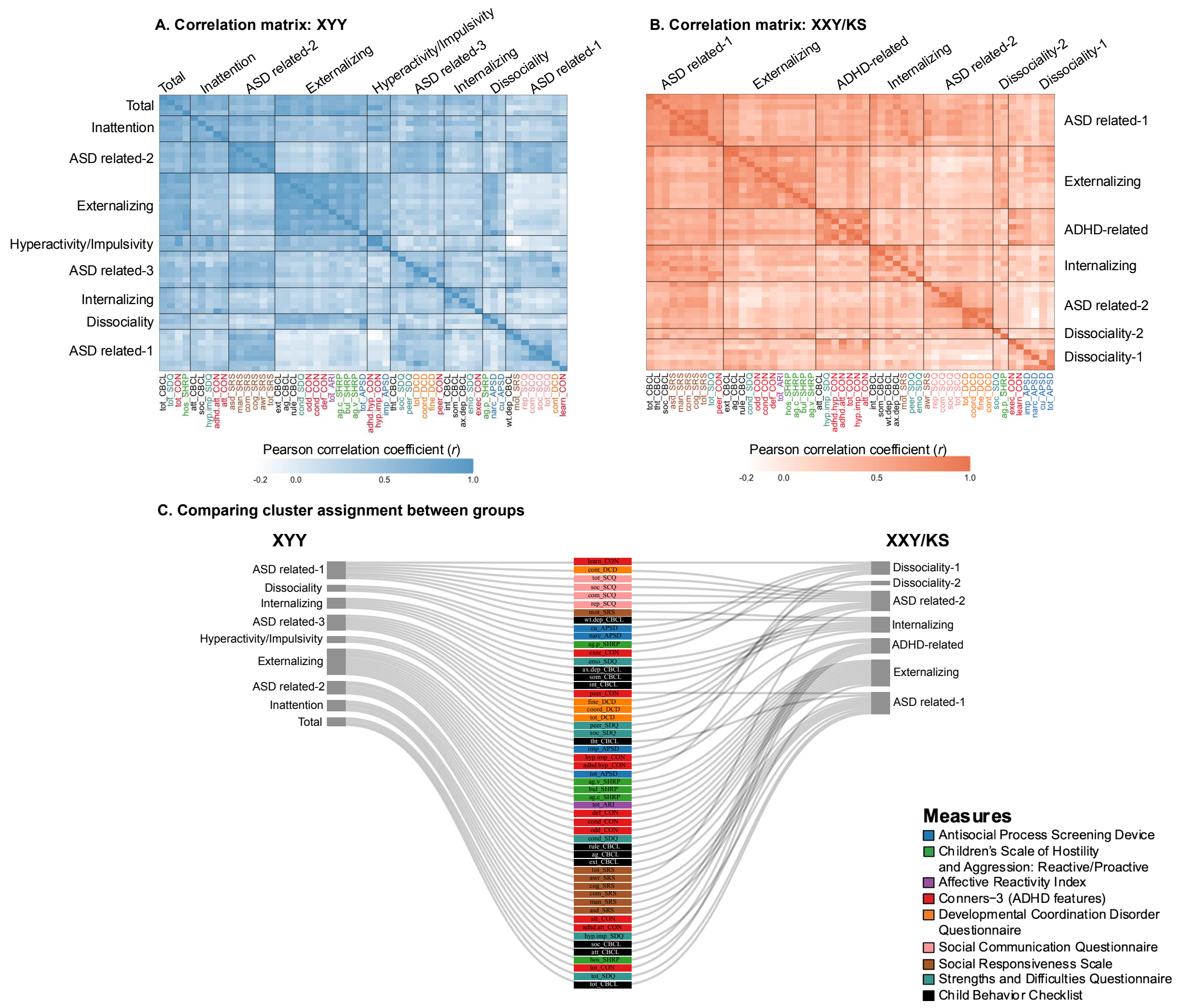


**Figure S2: Fine-grained symptom correlation and domain-level coupling for XXY/KS and XYY syndrome. (A)** Heatmap depicting the weighted stochastic block modeling (WSBM) solution of nine clusters (outlined in black) for the Pearson Correlation Coefficients between scales for XYY syndrome. **(B)** Heatmap depicting the weighted stochastic block modeling (WSBM) solution of seven clusters (outlined in black) for the Pearson Correlation Coefficients between scales for XXY/KS. The hue intensity depicts the magnitude of group differences in correlation between each unique pair of scales. **(C)** Sankey diagram depicting the cluster membership of the 53 scales in XYY syndrome versus XXY/KS. Scale names are color-coded to the instrument (see **Table S1**).

### Table S1

| Intrument (Abbreviation) | Subscale | Variable Name | *n* (XXY/KS, XY) | *n* (XYY, XY) | Mean age (XXY/KS, XY) | Mean age (XYY, XY) | Global coupling (XXY/KS) | Global coupling (XYY) | Deming residual | Divergence score | *p*-value against null distribution | Absolute delta effect size (XXY/KS - XYY) |
|---|---|---|---|---|---|---|---|---|---|---|---|---|
| **Antisocial Process Screening Device (APSD)** | Total | tot_APSD | 101, 73 | 58, 55 | 16.21, 17.17 | 13.24, 13.78 | 0.35 | 0.48 | 0.11 | 0.23 | 0.208 | 0.72 |
| | Callous/Unemotional | cu_APSD | 101, 73 | 58, 55 | 16.21, 17.17 | 13.24, 13.78 | 0.29 | 0.28 | -0.03 | 0.54 | 0.453 | 1.07 |
| | Narcissism | narc_APSD | 101, 73 | 58, 55 | 16.21, 17.17 | 13.24, 13.78 | 0.17 | 0.36 | 0.18 | 0.25 | 0.308 | 0.58 |
| | Impulsivity | imp_APSD | 101, 73 | 58, 55 | 16.21, 17.17 | 13.24, 13.78 | 0.30 | 0.40 | 0.07 | 0.18 | 0.628 | 0.25 |
| **Children's Scale of Hostility and Aggression (SHRP)** | Verbal aggression | ag.v_SHRP | 100, 38 | 54, 45 | 16.16, 12.46 | 12.13, 11.46 | 0.42 | 0.54 | 0.09 | 0.16 | 0.474 | 1.28 |
| | Bullying | bul_SHRP | 100, 38 | 54, 45 | 16.16, 12.46 | 12.13, 11.46 | 0.42 | 0.50 | 0.05 | 0.29 | 0.316 | 1.34 |
| | Covert aggression | ag.c_SHRP | 100, 38 | 54, 45 | 16.16, 12.46 | 12.13, 11.46 | 0.45 | 0.56 | 0.07 | 0.15 | 0.692 | 1.07 |
| | Hostility | hos_SHRP | 100, 38 | 54, 45 | 16.16, 12.46 | 12.13, 11.46 | 0.44 | 0.60 | 0.14 | 0.20 | 0.617 | 1.95 |
| | Physical aggression | ag.p_SHRP | 100, 38 | 54, 45 | 16.16, 12.46 | 12.13, 11.46 | 0.31 | 0.38 | 0.04 | 0.67 | **0.021*** | 0.74 |
| **Affective Reactivity Index (ARI)** | Total | tot_ARI | 101, 38 | 59, 46 | 16.13, 12.46 | 13.14, 12.37 | 0.40 | 0.49 | 0.06 | 0.17 | 0.511 | 1.54 |
| **Conners-3 (CON)** | Inattention | att_CON | 63, 38 | 48, 44 | 13.15, 12.46 | 11.19, 12.2 | 0.43 | 0.55 | 0.09 | 0.20 | 0.502 | 1.05 |
| | Hyperactivity/Impulsivity | hyp.imp_CON | 63, 38 | 48, 44 | 13.15, 12.46 | 11.19, 12.2 | 0.47 | 0.46 | -0.04 | 0.09 | 0.540 | 1.34 |
| | Learning problems | learn_CON | 63, 38 | 48, 44 | 13.15, 12.46 | 11.19, 12.2 | 0.24 | 0.26 | 0.00 | 0.45 | 0.478 | 0.30 |
| | Executive functioning | exec_CON | 63, 38 | 48, 44 | 13.15, 12.46 | 11.19, 12.2 | 0.33 | 0.37 | 0.02 | 0.21 | 0.699 | 0.11 |
| | Defiance | def_CON | 63, 38 | 48, 44 | 13.15, 12.46 | 11.19, 12.2 | 0.54 | 0.51 | -0.06 | 0.12 | 0.418 | 1.76 |
| | Peer relations | peer_CON | 63, 38 | 48, 44 | 13.15, 12.46 | 11.19, 12.2 | 0.50 | 0.50 | -0.04 | 0.31 | 0.259 | 2.79 |
| | Total | tot_CON | 63, 38 | 48, 44 | 13.15, 12.46 | 11.19, 12.2 | 0.56 | 0.62 | 0.02 | 0.13 | 0.804 | 1.09 |
| | ADHD inattentive subtype | adhd.att_CON | 63, 38 | 48, 44 | 13.15, 12.46 | 11.19, 12.2 | 0.43 | 0.52 | 0.06 | 0.10 | 0.991 | 0.80 |
| | ADHD hyperactive subtype | adhd.hyp_CON | 63, 38 | 48, 44 | 13.15, 12.46 | 11.19, 12.2 | 0.44 | 0.42 | -0.05 | 0.10 | 0.498 | 1.07 |
| | Conduct disorder | cond_CON | 63, 38 | 48, 44 | 13.15, 12.46 | 11.19, 12.2 | 0.50 | 0.43 | -0.11 | 0.25 | **0.047*** | 1.20 |
| | Oppositional defiant disorder | odd_CON | 63, 38 | 48, 44 | 13.15, 12.46 | 11.19, 12.2 | 0.44 | 0.56 | 0.09 | 0.07 | 0.977 | 1.47 |
| **Developmental Coordination Disorders Questionnaire (DCD; inverted)** | Control | cont_DCD | 100, 38 | 62, 51 | 16.16, 12.46 | 12.77, 11.7 | 0.37 | 0.32 | -0.08 | 0.18 | 0.337 | 1.66 |
| | Fine Motor | fine_DCD | 100, 38 | 62, 51 | 16.16, 12.46 | 12.77, 11.7 | 0.29 | 0.34 | 0.04 | 0.35 | 0.134 | 0.91 |
| | Coordination | coord_DCD | 100, 38 | 62, 51 | 16.16, 12.46 | 12.77, 11.7 | 0.37 | 0.47 | 0.07 | 0.25 | 0.087 | 0.72 |
| | Total | tot_DCD | 100, 38 | 62, 51 | 16.16, 12.46 | 12.77, 11.7 | 0.41 | 0.49 | 0.05 | 0.17 | 0.069 | 1.41 |
| **Social Communication Questionnaire (SCQ)** | Total | tot_SCQ | 100, 37 | 62, 50 | 16.16, 12.38 | 12.77, 11.62 | 0.37 | 0.38 | -0.02 | 0.09 | 0.719 | 3.04 |
| | Reciprocal social interaction | soc_SCQ | 100, 37 | 62, 50 | 16.16, 12.38 | 12.77, 11.62 | 0.33 | 0.31 | -0.04 | 0.09 | 0.864 | 3.91 |
| | Communication | com_SCQ | 100, 37 | 62, 50 | 16.16, 12.38 | 12.77, 11.62 | 0.26 | 0.33 | 0.05 | 0.11 | 0.873 | 1.18 |
| | Repetitive Behavior | rep_SCQ | 100, 37 | 62, 50 | 16.16, 12.38 | 12.77, 11.62 | 0.29 | 0.25 | -0.07 | 0.29 | 0.206 | 1.75 |
| **Strength and Difficulties Questionnaire (SDQ)** | Emotional symptoms | emo_SDQ | 101, 37 | 50, 51 | 16.13, 12.38 | 10.72, 11.7 | 0.34 | 0.33 | -0.03 | 0.19 | 0.897 | 0.67 |
| | Conduct problems | cond_SDQ | 101, 37 | 50, 51 | 16.13, 12.38 | 10.72, 11.7 | 0.41 | 0.51 | 0.07 | 0.15 | 0.133 | 0.35 |
| | Hyperactivity/Innatention | hyp.imp_SDQ | 101, 37 | 50, 51 | 16.13, 12.38 | 10.72, 11.7 | 0.42 | 0.49 | 0.04 | 0.18 | 0.281 | 1.15 |
| | Peer relationship problems | peer_SDQ | 101, 37 | 50, 51 | 16.13, 12.38 | 10.72, 11.7 | 0.36 | 0.43 | 0.04 | 0.27 | 0.334 | 2.14 |
| | Prosocial behavior (inverted) | soc_SDQ | 101, 37 | 50, 51 | 16.13, 12.38 | 10.72, 11.7 | 0.34 | 0.38 | 0.02 | 0.69 | 0.081 | 0.83 |
| | Total difficulties | tot_SDQ | 101, 37 | 50, 51 | 16.13, 12.38 | 10.72, 11.7 | 0.57 | 0.67 | 0.07 | 0.26 | 0.267 | 1.25 |
| **Social Responsiveness Scale (SRS)** | Total score | tot_SRS | 101, 38 | 62, 51 | 16.13, 12.46 | 12.77, 11.7 | 0.65 | 0.64 | -0.05 | 0.09 | 0.133 | 1.93 |
| | Social awareness | awr_SRS | 101, 38 | 62, 51 | 16.13, 12.46 | 12.77, 11.7 | 0.39 | 0.49 | 0.08 | 0.14 | 0.751 | 0.98 |
| | Social cognition | cog_SRS | 101, 38 | 62, 51 | 16.13, 12.46 | 12.77, 11.7 | 0.55 | 0.56 | -0.02 | 0.13 | 0.810 | 0.75 |
| | Social communication | com_SRS | 101, 38 | 62, 51 | 16.13, 12.46 | 12.77, 11.7 | 0.61 | 0.61 | -0.03 | 0.12 | 0.143 | 2.09 |
| | Social motivation | mot_SRS | 101, 38 | 62, 51 | 16.13, 12.46 | 12.77, 11.7 | 0.39 | 0.31 | -0.11 | 0.28 | **0.026*** | 0.76 |
| | Restricted interests/repetitive behavior | man_SRS | 101, 38 | 62, 51 | 16.13, 12.46 | 12.77, 11.7 | 0.55 | 0.51 | -0.08 | 0.18 | 0.442 | 3.73 |
| | ASD social communication/interaction | asd_SRS | 101, 38 | 62, 51 | 16.13, 12.46 | 12.77, 11.7 | 0.63 | 0.64 | -0.03 | 0.09 | 0.132 | 1.59 |
| **Child Behavior Checklist (CBCL)** | Anxious/Depressed | ax.dep_CBCL | 61, 37 | 47, 46 | 12.88, 12.6 | 11.04, 12.37 | 0.45 | 0.30 | -0.17 | 0.25 | 0.578 | 0.27 |
| | Withdrawn/Depressed | wt.dep_CBCL | 61, 37 | 47, 46 | 12.88, 12.6 | 11.04, 12.37 | 0.43 | 0.27 | -0.19 | 0.37 | 0.231 | 1.96 |
| | Somatic complaints | som_CBCL | 61, 37 | 47, 46 | 12.88, 12.6 | 11.04, 12.37 | 0.31 | 0.28 | -0.06 | 0.49 | 0.097 | 1.34 |
| | Social problems | soc_CBCL | 61, 37 | 47, 46 | 12.88, 12.6 | 11.04, 12.37 | 0.59 | 0.58 | -0.05 | 0.54 | **0.009*** | 4.06 |
| | Thought problems | tht_CBCL | 61, 37 | 47, 46 | 12.88, 12.6 | 11.04, 12.37 | 0.54 | 0.42 | -0.16 | 0.51 | 0.286 | 1.58 |
| | Attention problems | att_CBCL | 61, 37 | 47, 46 | 12.88, 12.6 | 11.04, 12.37 | 0.47 | 0.55 | 0.05 | 0.20 | 0.672 | 1.44 |
| | Rule breaking | rule_CBCL | 61, 37 | 47, 46 | 12.88, 12.6 | 11.04, 12.37 | 0.39 | 0.48 | 0.07 | 0.12 | 0.700 | 0.12 |
| | Aggressive behavior | ag_CBCL | 61, 37 | 47, 46 | 12.88, 12.6 | 11.04, 12.37 | 0.58 | 0.59 | -0.02 | 0.13 | 0.129 | 0.99 |
| | Internalizing problems | int_CBCL | 61, 37 | 47, 46 | 12.88, 12.6 | 11.04, 12.37 | 0.51 | 0.39 | -0.15 | 0.28 | 0.279 | 1.64 |
| | Externalizing problems | ext_CBCL | 61, 37 | 47, 46 | 12.88, 12.6 | 11.04, 12.37 | 0.59 | 0.61 | -0.01 | 0.09 | 0.463 | 0.92 |
| | Total problems | tot_CBCL | 61, 37 | 47, 46 | 12.88, 12.6 | 11.04, 12.37 | 0.70 | 0.70 | -0.04 | 0.36 | 0.062 | 2.00 |

**Table S1: Supplemental information from all analyses**. Including: instrument abbreviation, subscales, and variable names; sample size per instrument in each group (XXY/KS, XYY, and matched XY groups); mean age of respondents per instrument and group; global coupling (scale degree) scores per SCA group and their Deming residual; divergence score per scale and its *p*-value resulting from permutation against the null distribution; and the absolute difference in effect size between groups (XXY/KS - XYY). Bolding and asterisk (*) denotes nominal statistical significance ($p < .05$). Bolding and double asterisk (**) denotes statistical significance after Bonferroni correction for multiple comparisons ($p < .05/53$, or $p < .0009$).
